# Germline intergenic duplications at Xq26.1 underlie Bazex-Dupré-Christol syndrome, an inherited basal cell carcinoma susceptibility condition

**DOI:** 10.1101/2022.02.12.22270762

**Authors:** Yanshan Liu, Siddharth Banka, Yingzhi Huang, Jonathan Hardman-Smart, Derek Pye, Antonio Torrelo, Glenda M. Beaman, Marcelo G. Kazanietz, Martin J Baker, Carlo Ferrazzano, Chenfu Shi, Gisela Orozco, Stephen Eyre, Michel van Geel, Anette Bygum, Judith Fischer, Zosia Miedzybrodzka, Faris Abuzahra, Albert Rübben, Sara Cuvertino, Jamie M. Ellingford, Miriam J. Smith, D. Gareth Evans, Lizelotte J.M.T Weppner-Parren, Maurice A.M. van Steensel, Iskander H. Chaudhary, D. Chas Mangham, John T. Lear, Ralf Paus, Jorge Frank, William G. Newman, Xue Zhang

## Abstract

**Background:** Bazex-Dupré-Christol syndrome (BDCS; MIM301845) is a rare X-linked dominant genodermatosis characterized by follicular atrophoderma, congenital hypotrichosis and multiple basal cell carcinomas (BCCs). Previous studies have linked BDCS to an 11.4 Mb interval on chromosome Xq25-27.1. However, the genetic mechanism of BDCS remains an open question.

**Methods:** To investigate the genetic etiology of BDCS, we ascertained eight families with individuals affected with BDCS (F1-F8). Whole exome (F1 and F2) and genome sequencing (F3) were performed to identify putative disease-causing variants within the linkage region. Array-comparative genomic hybridization and quantitative PCR were used to explore copy number variations (CNV) in BDCS families, followed by long-range gap-PCR and Sanger sequencing to amplify duplication junction and define the precise head-tail junctions, respectively. Immunofluorescence was performed in hair follicles, BCCs and trichoepitheliomas from BDCS patients and sporadic BCCs to detect the expression of corresponding genes. The *ACTRT1* variant (p.Met183Asnfs*17), previously proposed to cause BDCS, was evaluated with allele frequency calculator.

**Results:** In eight BDCS families, we identified overlapping 18-135kb duplications (six inherited and two *de novo*) at Xq26.1, flanked by *ARHGAP36* and *IGSF1*. We detected ARHGAP36 expression near the control hair follicular stem cells compartment, and found increased ARHGAP36 levels in hair follicles in telogen, BCCs and trichoepitheliomas from patients with BDCS. ARHGAP36 was also detected in sporadic BCCs from individuals without BDCS. Our modelling showed the predicted *ACTRT1* variants maximum tolerated minor allele frequency in control populations to be orders of magnitude higher than expected for a high-penetrant ultra-rare disorder, suggesting loss-of-function of *ACTRT1* is unlikely to cause BDCS.

**Conclusions:** Our data support the pathogenicity of intergenic duplications at Xq26.1, most likely leading to dysregulation of *ARHGAP36*, establish BDCS as a genomic disorder, and provide a potential therapeutic target for both inherited and sporadic BCCs.

## Background

Bazex-Dupré-Christol syndrome (BDCS, also called Bazex syndrome or follicular atrophoderma and basal cell carcinomas; MIM 301845) is a rare X-linked disorder characterized by congenital hypotrichosis, follicular atrophoderma (seen as “ice pick” marks, usually on the dorsum of hands and feet) and susceptibility to develop basal cell nevi and basal cell carcinomas (BCCs). The nevi and BCCs generally occur on sun-exposed areas, including the head, neck and face from the second decade of life [1]. Other features reported in some individuals with BDCS include persistent milia, hyperpigmented macules, hypohidrosis, and trichoepitheliomas (benign tumors arising from basal cells in the hair follicles, which rarely transform to BCCs) [2–9].

Clinically, BDCS overlaps with basal cell nevus syndrome (Gorlin syndrome; MIM 109400), which predisposes to multiple BCCs and is caused by heterozygous germline variants in *PTCH1* [10, 11] or *SUFU* [12]. Both genes encode members of the hedgehog signaling pathway, and variants in them result in dysregulated overexpression of Gli transcription factors. BCC is the most common skin cancer [13], with somatic mutations in genes encoding key components of Hedgehog-Patched-Gli signalling often present in sporadic BCCs [14]. BCCs arise from hair follicle stem cells and/or other epithelial stem cells reprogrammed to a follicular differentiation [15, 16]. Of note, X-linked inheritance pattern is unusual for a cancer predisposition syndrome. The ‘two-hit model’ for familial cancers caused by loss of heterozygosity of the remaining functional allele in an individual with a germline loss-of-function variant is unlikely to occur for an X-linked tumor suppressor gene. Hence, we hypothesized that BDCS is caused by genetic variants resulting in altered Hedgehog-Patched signaling or -Gli activity in follicular stem cells via a mechanism that does not result in direct loss of function.

## Methods

### Patient ascertainment

The procedures followed were in accordance with the ethical standards of the responsible committee on human experimentation (institutional and national), and written informed consents to take part in the present study, as well as to have the results of this work published were obtained from the participants. We identified patients and families diagnosed with BDCS based on clinical history and examination.

### *Sanger* sequencing

A single primer pair (ACTRT1-F: TAGGTATGATTTGCTTTCCTTGGC, ACTRT1-R-R: CAACCTAAAGATTCATGACATGACTC) was designed to amplify the full length of ACTRT1, which encompassed the single coding exon and its 5 ’ and 3 ’ untranslated regions (UTR). At least one affected individual from each family underwent Sanger sequencing.

All coding exons of *ARHGAP36* (exons 2-12) were amplified in the panel of lymphocyte DNA samples from individuals with clinical diagnoses of Gorlin syndrome without causal variants in *PTCH1* or *SUFU* and underwent Sanger sequencing on an ABI3730xl DNA Analyser (Life Technologies, Paisley, UK).

### Exome and Genome sequencing

Whole exome sequencing was performed by using SureSelect kits (Human All Exon 50Mb for Families 1 and 2 and Human All Exon v.5 for Family 3) (both Agilent, Santa Clara, CA, USA) were used for whole exome sequencing. Paired-end sequencing (~100bp) was performed on Illumina HiSeq2000 (Family 1 and 2) or HiSeq2500 platforms (Family 3). For the first two families, a minimum of 4.3 Gb of high-quality mappable data was generated, yielding a mean depth of coverage of 40-fold and 84% of target bases sequenced at 10x coverage. A minimum of 4.5 Gb of sequence was generated, yielding a mean depth of coverage of 80-fold and 95% of target bases sequenced at 20x coverage. The sequence data were mapped to the human reference genome (hg19) using Burrows Wheeler Aligner [17]. Variant calling was performed using the GenomeAnalysisToolKit-v2.4.7. (GATK) software [18]. Genome sequence data was generated by Complete Genomics (Mountain View, California, USA) as described previously [19]. Bioinformatics (alignment to the hg19 reference genome, local de novo assembly and variant calling) was performed using version 2.5 of the Complete Genomics pipeline [20].

### Array-comparative genomic hybridisation (a-CGH)

A-CGH was performed using Affymetrix SNP6.0 microarray as described previously [21].

### Real-time quantitative PCR (qPCR)

To validate the genomic duplications detected by aCGH and determine the boundaries of duplications in the three families and other families, qPCR was performed as previously described [22]. For families 1, 2 and 7, the primers for qPCR were designed according to aCGH results. For the other families, primer pair XQM located in the middle of the shared duplicated region (chrX:130348186-130348315) was designed. The quantification of the target regions was normalized to an assay from chromosome 21. The relative copy number (RCN) was determined with the comparative ΔΔCT method, using DNA from a normal male as the calibrator. All assays were repeated three times. A ~2-fold RCN indicated duplication in samples from males and ~3-fold RCN for samples from females. Primers used for each family were listed in Supp Table 3.

### Haplotype analysis

Four common single nucleotide polymorphisms (SNPs) (rs62603806, rs4240127, rs5932866, rs12559533) within the duplicated region were amplified with primer pair (SNP-F: GCACAGATGATTATGTCTGTTCC, SNP-R: CTGTCCCTACTTAGTAAATCGAG) and Sanger sequenced to generate haplotypes in the male patients of F1, F3, and F8.

### Long-range Gap-PCR

A series of qPCR primers was designed to walk through to refine the duplication boundaries. Once the boundaries were estimated by qPCR, the forward primer from the centromeric side of the duplication region that was nearest to the determined boundaries, as well as the reverse primer from the telomeric side were chosen to perform long-range gap-PCR in order to amplify the duplication junctions. F3, F5 and F8 used the same primers pair as F1. Sanger sequencing was conducted to define the precise breakpoints. Primers used for each family were listed in Supp Table 3.

### Population screening

Primer pair XQM, as described above, was used to determine the frequency of duplications at the disease associated locus by qPCR. The criteria defining duplication was the same as above.

### Immunohistochemistry

Immunohistochemistry was performed on 7um frozen sections by fixation in chilled paraformaldehyde (4% v/v) for 10 minutes. After washing with phosphate buffered saline (PBS), sections were permeabilised with PBS containing 0.1% Triton X100 for ten minutes. Primary antibodies were incubated over night at 4°C at a concentration of 1:200 (ARHGAP36, HPA002064, Sigma, Dorset, UK), 1:20 (IGSH, HPA035582, Sigma) or 1:100 (ACTRT1, HPA003119, Sigma). For dual stains cytokeratin 15 (Abcam, ab80522, Cambridge, UK), at a concentration of 1:500 was added and incubated overnight.

Antibodies were detected by incubating with an Alexafluor goat anti mouse/rabbit secondary (Sigma) antibody at a concentration of 1:200 for 45 minutes. Sections were counterstained with DAPI.

For tumor tissue, paraffin sections were de-waxed using xylene and rehydrated using decreasing concentrations of ethanol (100%-50%). Antigen retrieval was performed by boiling in 10mM citrate buffer (pH 6) for 20 minutes. Following this the protocol as above was followed with P63 (Abcam-ab735) at a concentration of 1:100.

### Allele frequency modelling

Allele frequency modelling of *ACTRT1* was performed using the allele frequency calculator from http://cardiodb.org/allelefrequencyapp [23].The maximum tolerated reference allele count (0.95 CI) for the ACTRT1 NM_138289.3:c.547dup (p.(Met183Asnfs*17)) variant was calculated in 204,684 in alleles (https://gnomad.broadinstitute.org/variant/X-127185638-A-AT?dataset=gnomad_r2_1). The adjustable parameters were – estimated population prevalence of BDCS (1 in106) (Orpha.net); allelic heterogeneity of 33% (BDCS in 2 out of 6 families in Bal et al was attributed to this variant); and penetrance of 100% (based on description of families in the literature. Next, to account for potential inaccuracies in previous estimates, we modelled the maximum tolerated reference allele counts with various combinations of prevalence or penetrance values.

## Results

### BDCS is caused by small tandem intergenic duplications at chromosome Xq26.1

Previous mapping has linked BDCS to an 11.4 Mb interval on chromosome Xq25-27.1 [24, 25]. To further investigate the genetic etiology of BDCS, we ascertained eight families (F1-8) with individuals affected with BDCS (Fig. 1), including five previously published families (F1 [25], F4 [26], F5 [27], F7 [3], and F8 [28]). Whole exome (F1 and F2) or genome (F3) sequencing did not identify putative disease-causing variants within the Xq26.1 locus previously linked to BDCS [25]. Array-comparative genomic hybridization in at least one affected individual from F1, F2, F3, and F7 revealed small intergenic Xq26.1 gains of varying sizes (Fig. 2A) [25]. Further, qPCR assays in affected individuals from the other four families (F4, F5, F6, and F8) were consistent with gains at this locus and confirmed that the gains segregated with BDCS in the multiplex families or were *de novo* in the two simplex cases (Fig. 2B). Long-range gap-PCR to amplify the duplication junctions followed by Sanger sequencing defined the head-tail junctions for F2, F4, F6 and F7 and confirmed these gains as tandem duplications (Fig. 2C). The duplications in F1, F3, F5, and F8 could not be differentiated further, likely due to homologous L1 elements (Fig. 2D). In the three multiplex families (F1, F3 and F8) with seemingly identically sized duplications, SNP haplotype mapping proved their likely independent origin (Fig. 2E). No gains were identified by qPCR in 215 unrelated European controls (139 females and 76 males). The largest gain was detected in F2 (135kb) and the 18kb gain in F6 defined the smallest shared overlapping region (hg19, chrX:130,341,750-130,360,310) (Fig. 2D).

**Fig. 1.**
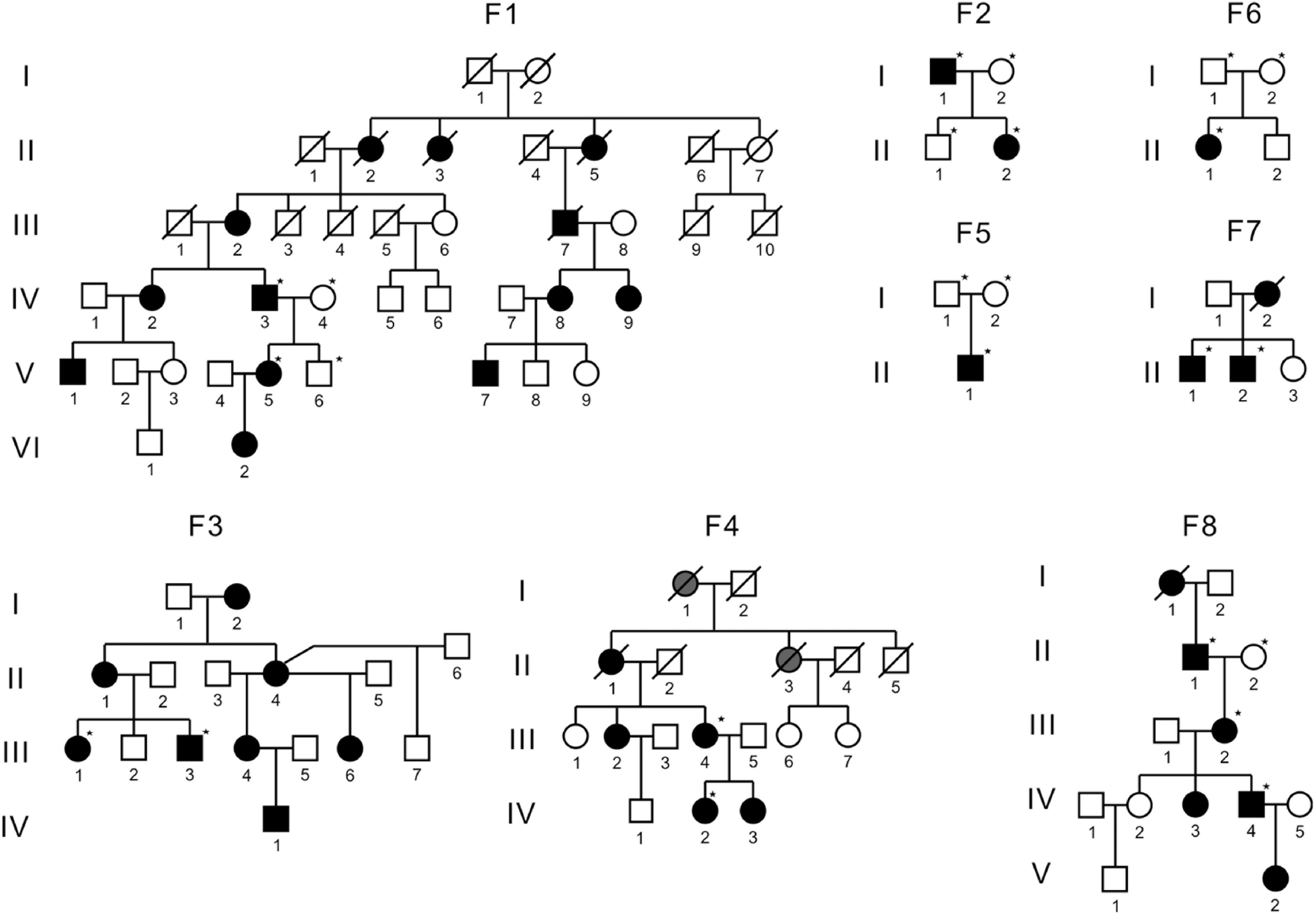
Individuals with BDCS and pedigrees of families. Pedigrees of all families included in this study are shown. Standard symbols have been used to denote sex and affected status. * indicates individuals from whom DNA samples were available for analysis. At least one affected individual from each family underwent *ACTRT1* Sanger sequencing including its full coding region and the 5’ and 3’ untranslated regions. Figures of BDCS patients from current study were available from the corresponding authors on reasonable request.

**Fig. 2.**
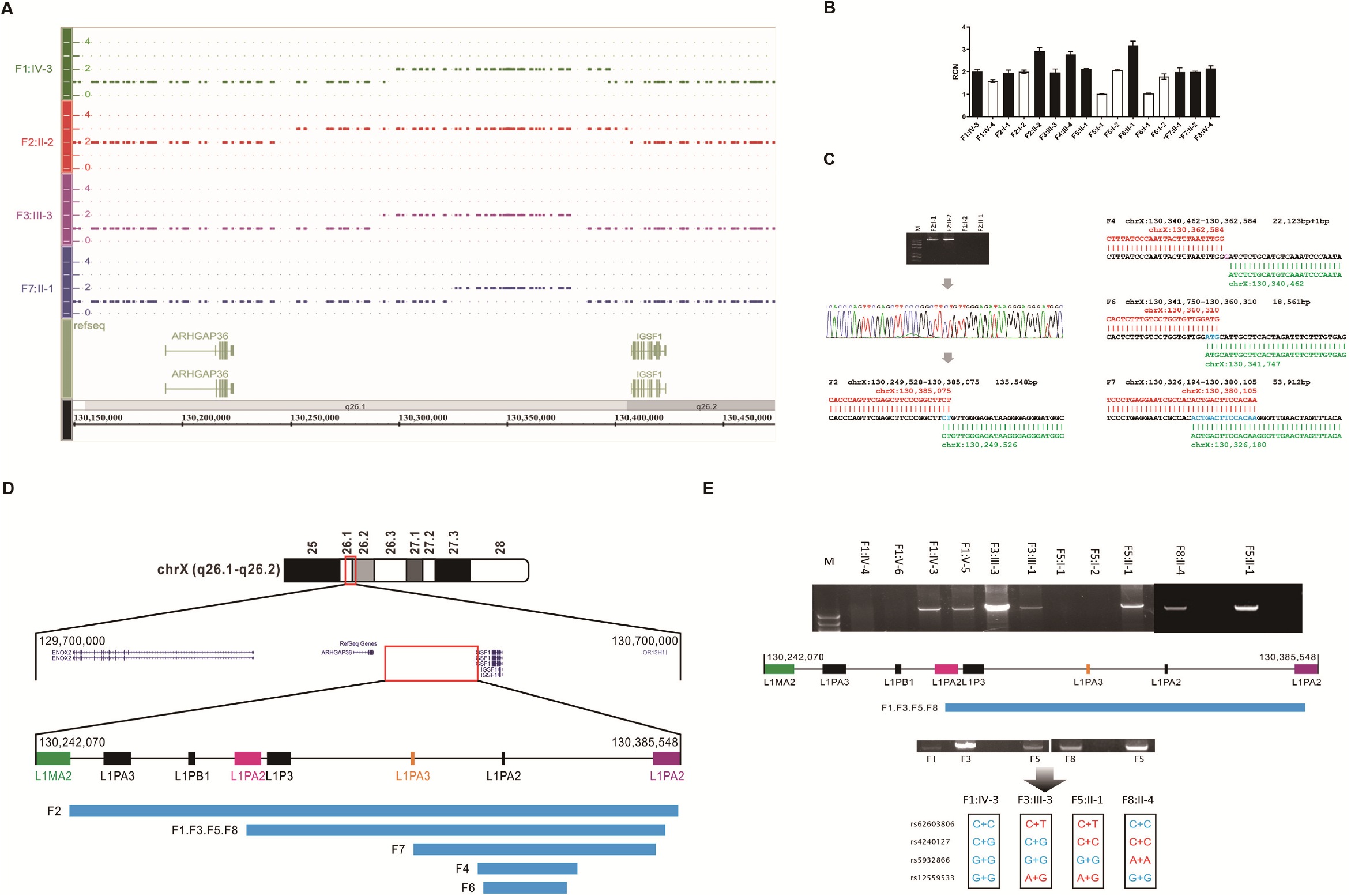
Small intergenic duplications of chromosome Xq26.1 in individuals with BDCS. (A) Array comparative genomic hybridisation in four individuals with BDCS, demonstrating intergenic copy number gains at the Xq26.1. The top four panels show the corresponding copy number of F1:IV-3, F2:II-2, F3:III-3 and F7:II-1 between chrX: 130,150,000-130,450,000 (hg19), respectively. The bottom panel shows the flanking RefSeq protein coding genes. (B) qPCR of an amplicon at the duplicated locus demonstrating normal copy number (white bar) in the unaffected mothers of individuals F5:II-1 and F6:II-1 consistent with de novo origin of the duplications in these affected (black bar) individuals. In other families the presence of the duplications segregated with the phenotype. *indicates different primer pair was used in F7 compared with other families. (C) The specific breakpoints for the duplications defined in affected individuals from families F2, F4, F6 and F7. (D) A cartoon of the duplications at Xq26.1-26.2 from the eight families with BDCS, with individual F6:II-1 defining the boundaries of the critical interval. (E) Long-range gap PCR in families’ members of F1, F3, F5 and F8 had seemingly identically sized duplications (top). Haplotype analysis with four common single nucleotide polymorphisms (SNPs) (rs62603806, rs4240127, rs5932866, rs12559533) within the duplicated region demonstrates independent origins of F1, F3 and F8 (down).

No similar sized exclusively intergenic gains overlapping with the shared duplicated region defined by the eight families were detected in control populations (Fig. S1). One entirely intergenic gain (nsv517789; chrX:130,234,700-130,340,623) of ~100kb, in an individual with no known phenotype was noted, but it did not overlap the shared duplicated region as defined by our study (Fig. S1) [29]. Another gain of >380kb (nsv528179; chrX:130,300,617-130,680,930) overlapping the shared BDCS duplicated region, in an individual with no known phenotype was noted, but it extended ~295kb beyond the most telomeric boundary of the BDCS associated duplications and encompasses *IGSF1* (Fig. S1). Other larger chromosome X duplications encompassing the region have also been reported in individuals without BDCS [29].

### The Xq26.1 duplications in BDCS likely dysregulate ARHGAP36

None of the BDCS duplications encompass protein-coding genes, suggesting dysregulation of flanking genes as the likely disease-associated mechanism (Fig. 2D). BDCS is considered to be a disorder of the hair follicle [5]. We, therefore, performed immunofluorescence for the two flanking genes, ARHGAP36 and IGSF1 in control hair follicles. In anagen, IGSF1 was present in the terminally differentiated inner root sheath (IRS), but was absent from the actively proliferating hair matrix or bulge, the main reservoir of hair follicle epithelial stem cells (Fig. S3a). In telogen, there was no evidence of IGSF1 staining (data not shown). In contrast, ARHGAP36 was present in a small number of hair follicle cells in the outer root sheath (ORS) at the level of the stem cell bulge, in both anagen (Fig. 3A) and telogen (Fig. 3C). Notably, it is at the end of telogen that the stem cells located in the secondary hair germ and bulge regions are activated to resume hair growth [30]. There was no obvious difference regarding ARHGAP36 positive cell numbers in the ORS between hair follicles from healthy control and an individual with BDCS (F5:II-1) in anagen (Fig. 3A and 3B). In contrast, in telogen hair follicle from F5:II-1, there was a marked increase in the number of ARHGAP36 positive cells around the epithelial stem cell compartment (Fig. 3D), comparing with healthy control in telogen (Fig.3C) and the same patient in anagen (Fig. 3B). Moreover, immunofluorescence in histologically confirmed p63 positive BCC [31] from F4:III-4 showed strong staining for ARHGAP36 in a proportion of cells (Fig. 4A-B). Notably, the BCCs did not immunofluoresce for IGSF1 (Fig. S4a). We also detected striking ARHGAP36 staining of a trichoepithelioma from the same individual (Fig. 4C).

**Fig. 3.**
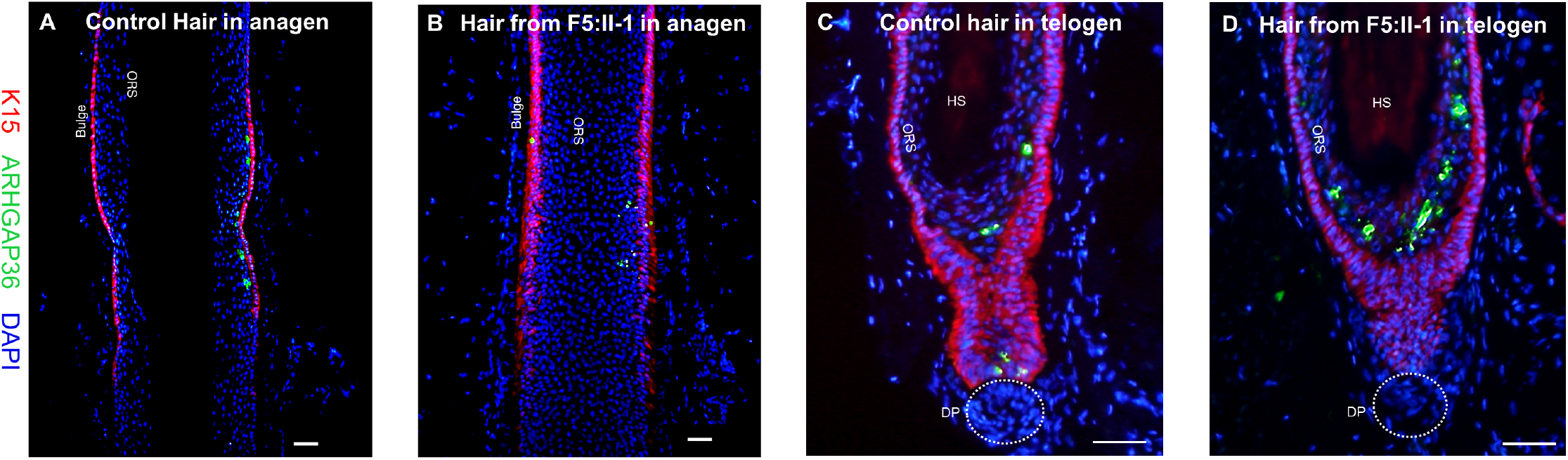
Immunofluorescence of hair follicles in anagen and telogen from normal healthy skin and an individual (F5:II-1) with BDCS for ARHGAP36. A small number of cells in the stem cell bulge region stain positively (green) for ARHGAP36 in the hair follicle from normal healthy skin (A) and the skin from F5:II-1 (B). (C) A normal hair follicle in telogen stained for ARHGAP36 (green) demonstrating a small number of positively stained cells in the outer root sheath adjacent to K15 positive bulge stem cells (red). (D) A hair follicle in telogen from F5:II-1 showing an increased number of positively stained cells. Red stain for keratin 15. (DP = dermal papilla; ORS = outer root sheath; HS = hair shaft). Scale bar 50μm.

**Fig. 4.**
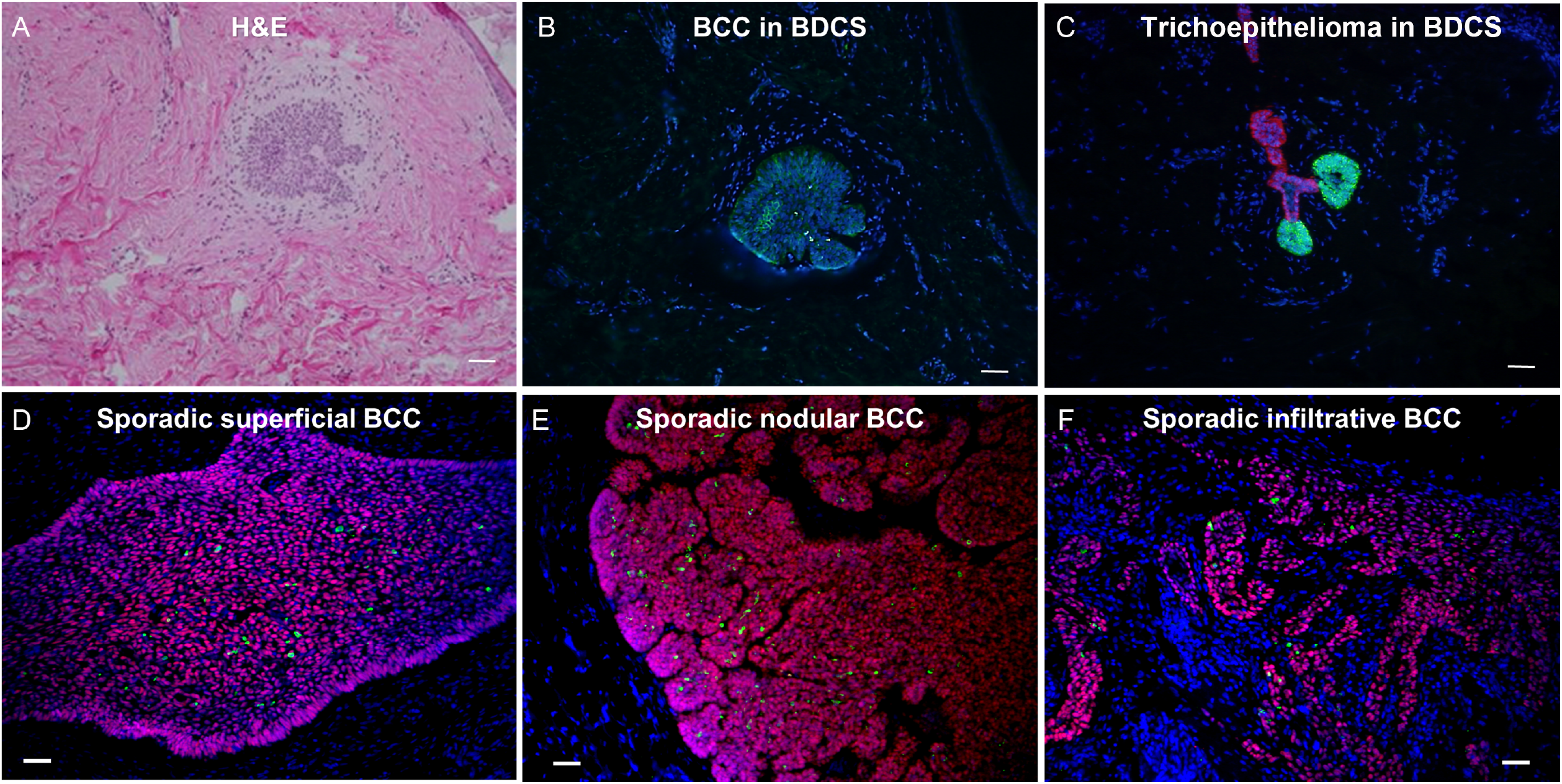
Immunofluorescence of ARHGAP36 in BDCS patient and sporadic BCCs. (A-C) Pathological study of tumors. Haematoxylin and eosin staining (A) and immunofluorescence (B) in a BCC from an individual with BDCS (F4:III-4) demonstrating staining (green) for ARHGAP36 in the tumor but not in the surrounding tissue. Strong staining of a trichoepithelioma (C) for ARHGAP36 and P63 (pink) in the same individual. Superficial (D), nodular (E) and infiltrative (F) sporadic BCCs from individuals without BDCS stained for ARHGAP36 (green) and and P63 (pink). Scale bar 50μm.

Next, to explore if ARHGAP36 could be relevant to sporadic BCCs, we determined the presence of ARHGAP36 in superficial (n=10), nodular (n=10), and infiltrative (n=10) sporadic BCCs. Similar to the BCCs in BDCS, ARHGAP36 was present in a small proportion of cells from all examined tumors (Fig. 4D-F), but was absent from all surrounding tissue.

### *ACTRT1* loss-of-function variants are unlikely to cause BDCS

Knudson’s two-hit hypothesis explains the mechanism of most inherited cancer syndromes, in which tumors occur following somatic loss of the only functional allele in an individual with a germline loss-of-function variant in a tumor suppressor gene [32]. Presence of a single X-chromosome in males and X-inactivation in females, make X-linked inherited cancer predisposition syndromes due to tumor gene suppression highly unlikely. Previously, Bal *et al* reported loss-of-function *ACTRT1* variants in BDCS [28]. They identified a frameshift *ACTRT1* NM_138289.3: c.547dup (p.Met183Asnfs*17) variant (rs771087307), in two families with BDCS [28]. Variants in conserved non-coding elements flanking *ACTRT1* were also proposed as pathogenic. Our modelling revealed that the predicted maximum tolerated minor allele frequency (MAF) for the p.Met183Asnfs*17 variant in population control data to be ~10^4^ times higher than expected and could be reconciled only if the previous prevalence and penetrance estimates were inaccurate by several orders of magnitude (Fig. 5). Furthermore, other putative loss-of-function variants in the single exon of *ACTRT1* have been reported in both males and females without BDCS (Tables S1-2). Sanger sequencing in at least one affected individual from F1-F7 did not identify any rare coding variants in *ACTRT1*. As expected, the p. Met183Asnfs*17 variant was present in the affected individuals (II-1 and III-1) in F8, as reported in the original association paper [28]. Importantly, immunofluorescent staining showed that ACTRT1 was absent from both the disease-relevant regions of the hair follicles and tumors from individuals with BDCS (Fig. S3b and S4b). These combined data provide evidence that *ACTRT1* loss-of-function variants are unlikely to cause BDCS.

**Fig. 5.**
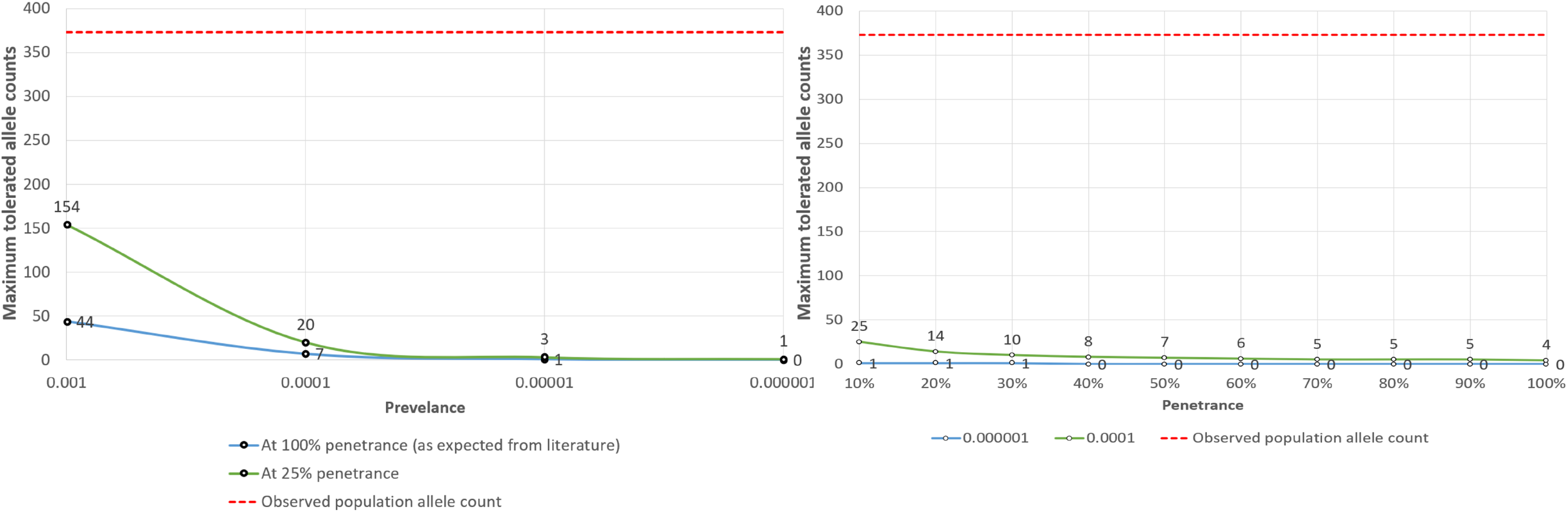
ACTRT1 loss-of-function variants are unlikely to cause BDCS. Modelling of maximum tolerated allele counts (MTAC) for the *ACTRT1* NM_138289.3:c.547dup (p.(Met183Asnfs*17)) variant is shown. The left and right panels show MTACs against different levels of possible prevalence and penetrance respectively. In both panels, the blue lines model MTACs at the prevalence (left panel) or penetrance (right panel) levels consistent with the existing literature. The green lines model MTACs at much lower hypothetical constraints. Allele count estimates are for 204,684 alleles in the reference population at 0.95 confidence interval, 33% allelic heterogeneity and genetic heterogeneity of 1. Note that the observed population allele count (=373) of the variant, shown in broken red line, is significantly higher than estimated maximum tolerated allele counts across all the modelled scenarios.

## DISCUSSION

We show that BDCS is caused by small intergenic tandem duplications at Xq26.1 that encompass a minimum shared overlapping region of ~18Kb (chrX:130,341,750-130,360,310) (Fig. 2). Identification of a BDCS duplication at this locus in a family previously reported to have a causal *ACTRT1* variant is indicative of the likely benign nature of *ACTRT1* variants in individuals with BDCS [28, 33]. Importantly, our data suggests that larger Xq26.1 duplications *encompassing* the flanking genes in addition to the minimum common shared region may not result in BDCS. Also, smaller ‘non-coding’ duplications at Xq26.1 that do not overlap the shared minimum overlapping region may not cause BDCS. These observations reflect the complexity of the genetic diagnosis of BDCS and assigning pathogenicity to Xq26.1 duplications in diagnostic labs will require careful consideration of the size and location of the copy number gains at this position. Future studies will be needed to validate our findings and to refine the minimum common overlapping region for BDCS duplications.

To the best of our knowledge, BDCS is the first example of an inherited cancer predisposition disease caused by germline CNVs that do not encompass any protein coding genes. To explore the effects of the intergenic BDCS duplications, we first turned our attention to the two flanking genes, as our preliminary Hi-C assay on dermal fibroblasts from health control and BDCS patients showed that the of BDCS duplications did not disrupt the topologically associated domain (TAD), which contained IGSF1 and ARHGAP36 (Fig. S5). *IGSF1*, the flanking telomeric gene, encodes member 1 of the immunoglobulin superfamily and loss-of-function variants in this gene cause an X-linked recessive syndrome of central hypothyroidism and testicular enlargement (MIM 300888) [34]. There is no known role of IGSF1 in Hedgehog-Patched-Gli pathway. *ARHGAP36*, the flanking centromeric gene, encodes a Smoothened (Smo)-independent positive regulator of the sonic hedgehog (shh) pathway and its expression is upregulated in medulloblastomas, a Hedgehog-Patched-Gli pathway-related tumor [35, 36]. Variants in *ARHGAP36* have not been associated with any inherited disorder. Hence, the known biological role makes ARHGAP36 an excellent candidate for BDCS pathology. In this context, our finding of ARHGAP36 expression is increased in telogen phase in follicles, in BCCs and trichoepithelioma from BDCS patients (Fig. 3D, 4B and 4C) makes it highly likely that ARHGAP36 dysregulation is the likely disease mechanism.

Interestingly, our results suggest that ARHGAP36 is also relevant to sporadic BCC pathology (Fig. 4D-F). Of note, ARHGAP36 did not co-localize with p63 (Fig. 4C-F) and the higher levels of ARHGAP36 in trichoepithelioma than in the BDCS-associated and sporadic BCCs suggests that dysregulation of ARHGAP36 may be an early step in the pathogenesis to BCC. ARHGAP36, therefore, could be an attractive therapeutic target for inhibition in individuals with both inherited and, the vastly more common, non-inherited forms of BCC.

ARHGAP36 is a member of the Rho GAP family of regulatory proteins, which deactivate Rho proteins. Rac1 (a RhoGTPase) is essential for hair follicle stem cell function [37, 38]. With pulldown assays, we demonstrated interaction between ARHGAP36 and RAC1 (Fig. S6). This preliminary finding potentially uncovers a previously unknown role of ARHGAP36 and may explain the non-cancerous phenotypes of BDCS, namely hypotrichosis. However, further confirmatory studies will be required to solve the mystery underlying *ARHGAP36* dysregulation and BDCS phenotypes.

## Conclusions

In summary, we have shown that small intergenic non-coding tandem duplications at Xq26.1 encompassing chrX:130,341,750-130,360,310 (hg19) cause BDCS. This is the first example of an inherited cancer predisposition disease caused by germline non-coding CNVs. We propose that the duplications result in the dysregulation of *ARHGAP36* that underlies BDCS pathology. Our findings reconcile the molecular mechanism of BDCS, a tumor-predisposition syndrome, with its X-linked inheritance pattern. We also suggest that ARHGAP36 is relevant to sporadic BCCs and a potential therapeutic target.

## Data Availability

All data produced in the present work have not been deposited in a public repository due to privacy and ethical restrictions, but are available from the corresponding authors on request.

## Authors’ contributions

XZ, JF and WGN conceived the project. XZ designed and coordinated the genetics study. WGN and SB designed and directed the histopathology study. YL, YH, MGK, MJB, GO, SE, JTL and RP conducted the experiments. YL, YH, JHS, DP, GMB, MGK, CF, CS, SC, JME, MJS, IHC, SB, AT, MvG, AB, JF, ZM, FA, AR, DGE, LJMTW-P, MAMvS, DCM, JF and WGN collected clinical data. SB, YL, YH, JF, WGN and XZ wrote the manuscript.

## Availability of data and materials

WES, WGS and array-comparative genomic hybridization datasets have not been deposited in a public repository due to privacy and ethical restrictions, but are available from the corresponding authors on reasonable request.

## Ethics approval and consent to participate

The procedures followed were in accordance with the ethical standards of the responsible committee on human experimentation (institutional and national), and written informed consents to take part in the present study, as well as to have the results of this work published were obtained from the participants.

