## supplemental figures and tables for "Germline intergenic duplications at Xq26.1 underlie Bazex-Dupré-Christol syndrome, an inherited basal cell carcinoma susceptibility condition"

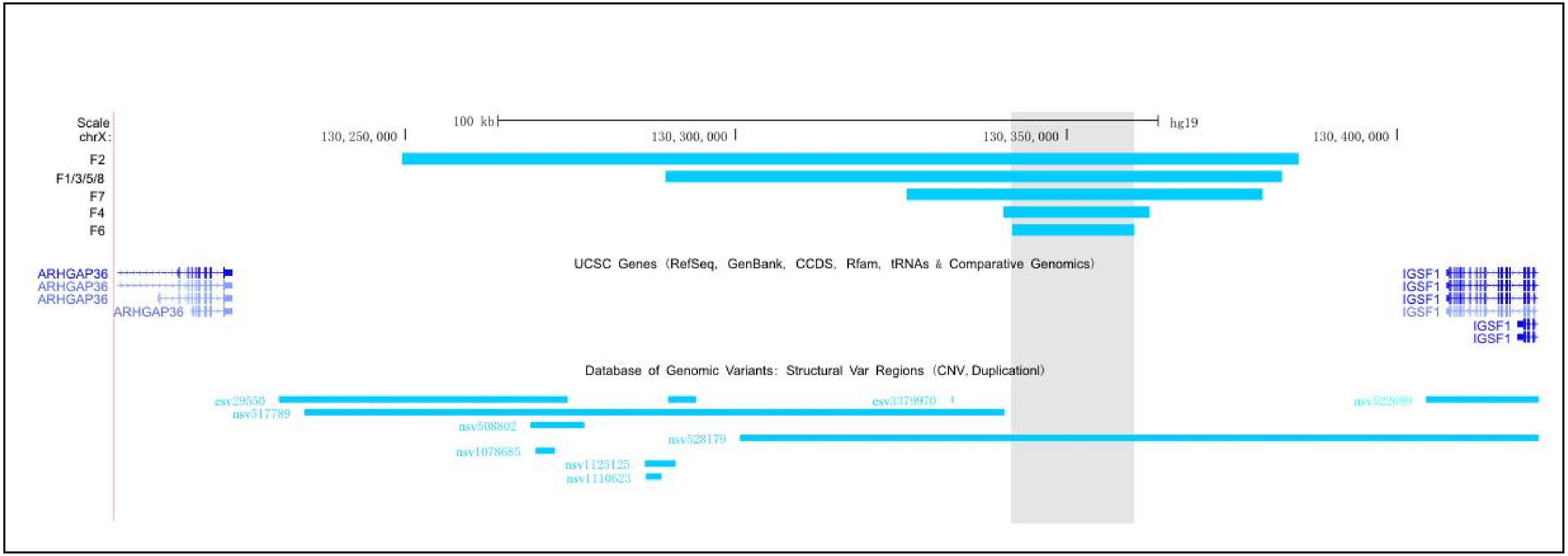

**Figure S1. No similar sized duplications were exclusively intergenic 2 and overlapped with the shared duplicated region defined by BDCS families.** Relative location of the long structure variation at Xq26.1-26.2 from the Database of Genomic Variants (https://www.ebi.ac.uk/dgva/) and BDCS associated duplications. The figure was generated and modified from UCSC genome browser ( https://genome.ucsc.edu/). Duplications from BDCS families were induced as custom track. Default colors were used to indicate different structure variations. Duplication (gain in size)was indicated in blue.

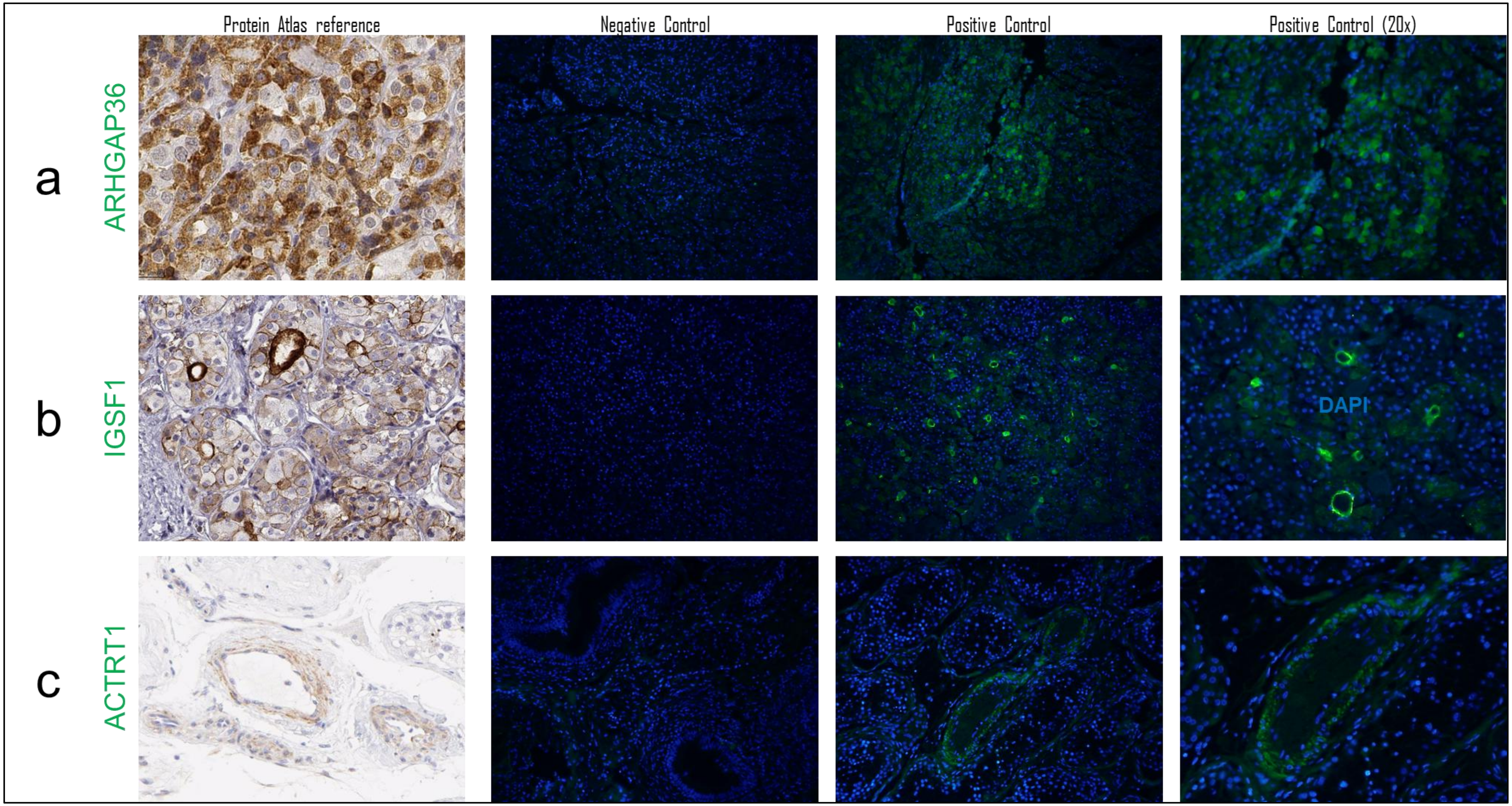

**Figure S2. Immunohistochemistry staining patterns of ARHGAP36, IGSF1 and ACTRT1.**Immnuohistochemistry staining figures from the Protein Atlas Reference (https://www.proteinatlas.org) in pituitary gland for a) ARHGAP36 and b) IGSF1 and testes for c) ACTRT1 were used as reference. Brown positive staining for the respective proteins. Negative (without antibody) controls for the three proteins in the respective tissue. Immunofluorescence for the three antibodies show staining patterns comparable to the reference data supporting the specificity of the antibodies. DAPI=blue. Green stain relates to the presence of a) ARHGAP36 b) IGSF1 and c) ACTRT1.

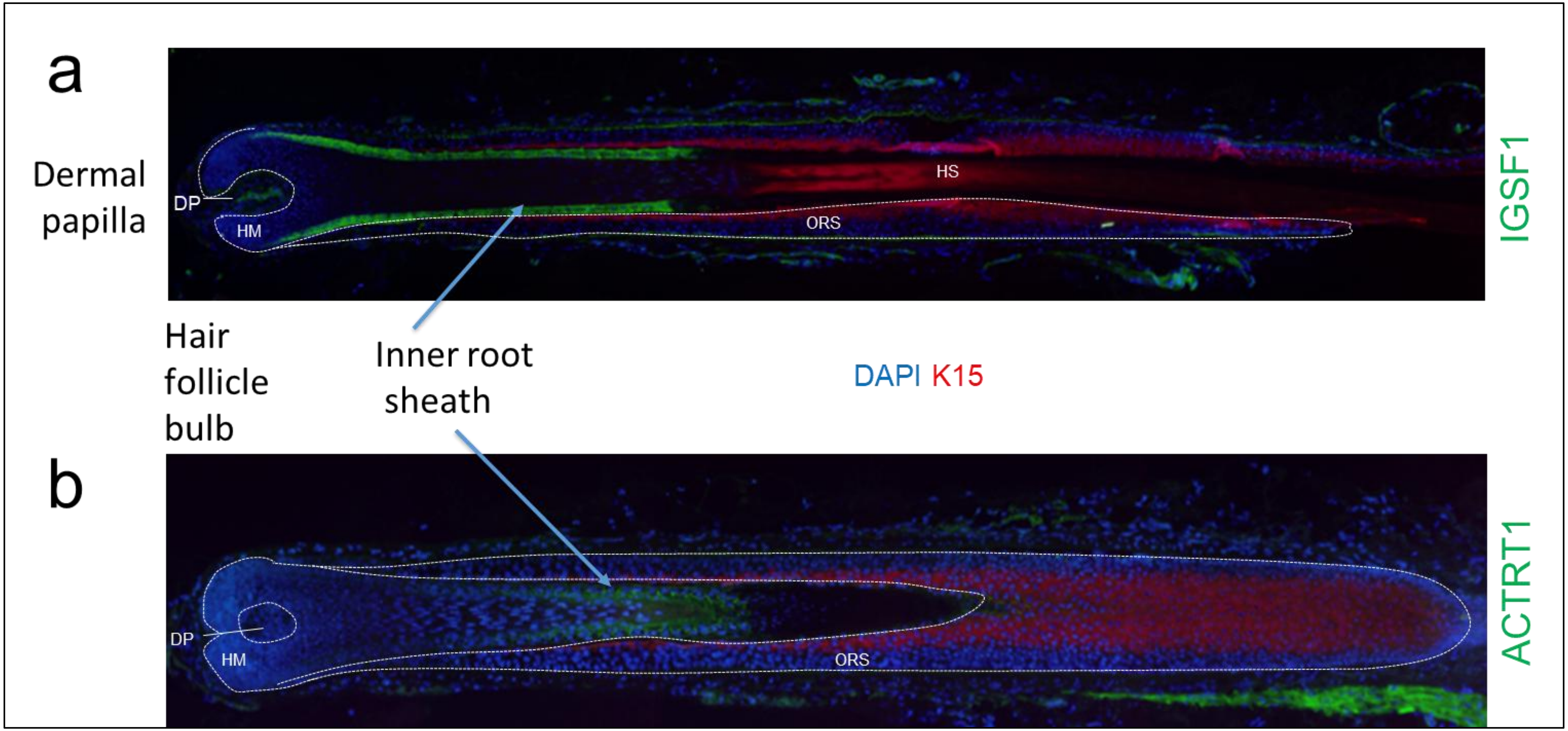

**Figure S3. Expression and localisation of IGSF1 and ACTRT1 in anagen in hair follicle from normal healthy skin.** Immunofluorescence of hair follicles in anagen from normal skin for IGSF1 and ACTRT1 (green) with staining of the inner root sheath marked by arrows. Note no staining for either IGSF1 or ACTRT1 at the stem cell bulge region. DAPI =blue and Keratin K15=red. K15 is a marker of epidermal stem cells. (DP = dermal papilla; ORS = outer root sheath; HS = hair shaft)

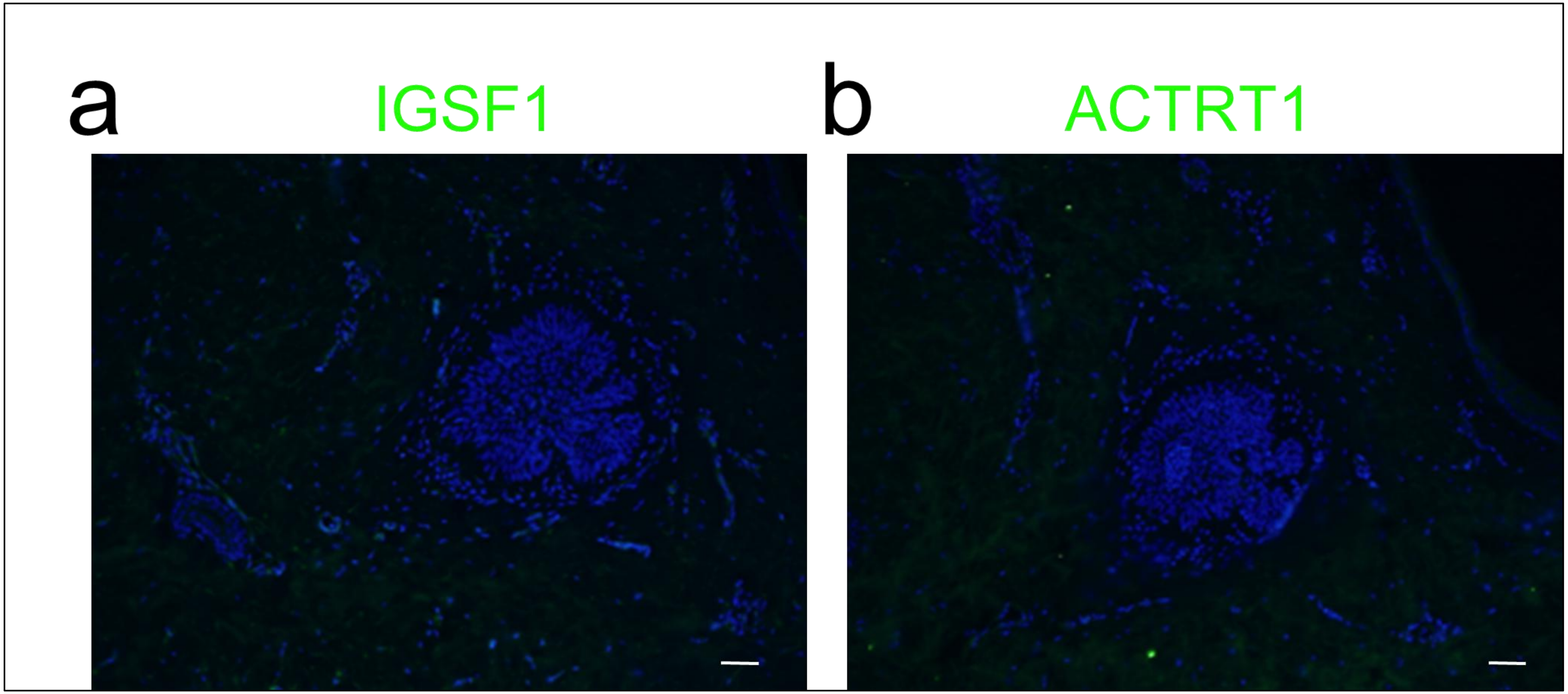

**Figure S4. Immunofluorescence of BCC tumors from an individual with BDCS (F5:II-1).** Absence of immunofluorescence for (a) IGSF1 and (b) ACTRT1 respectively in a BCC.

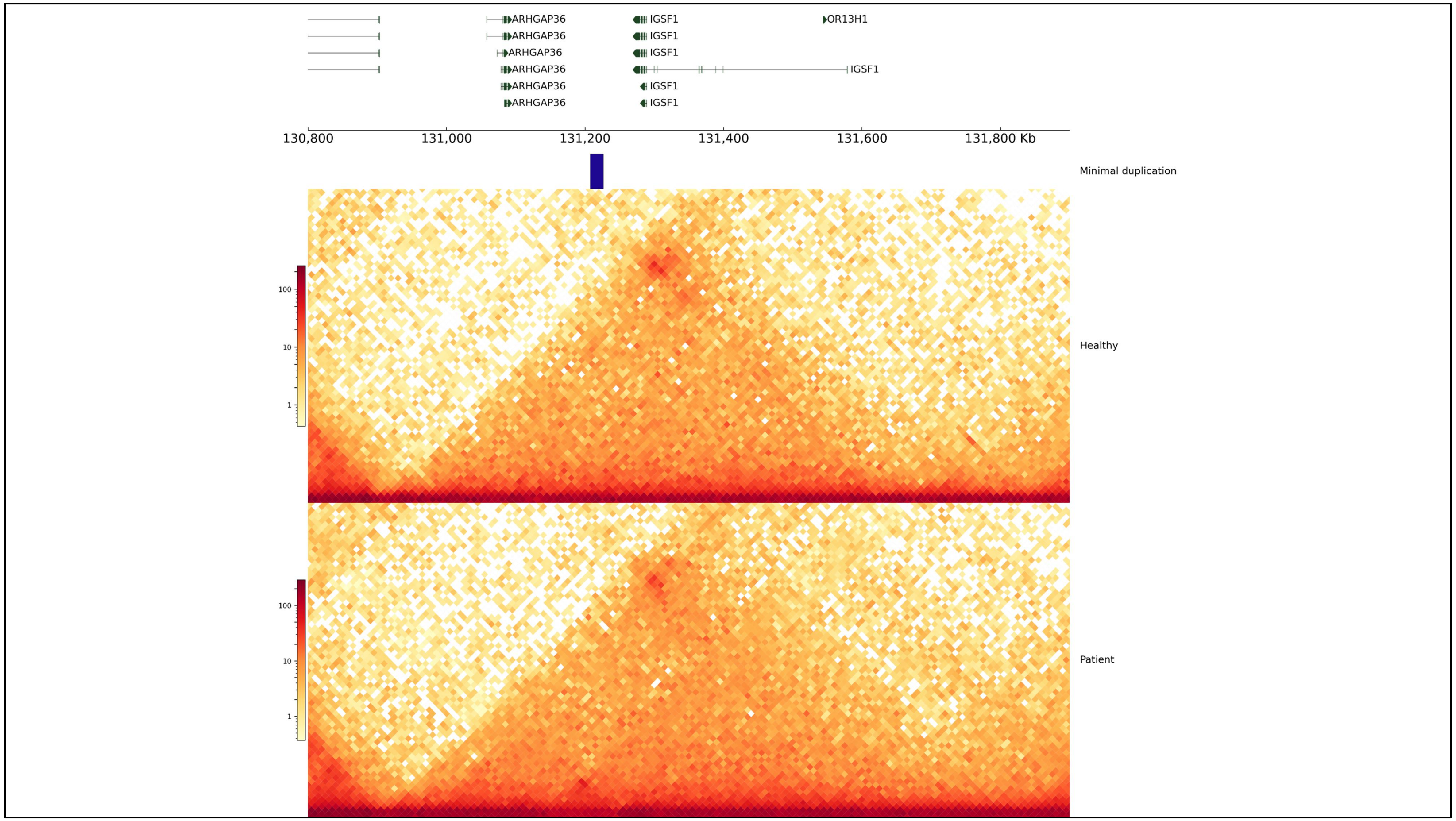

**Figure S5. All the duplication regions detected in BDCS patients were located within a single topologically associated domain (TAD) containing ARHGAP36, IGSF1 and OR13H1.**

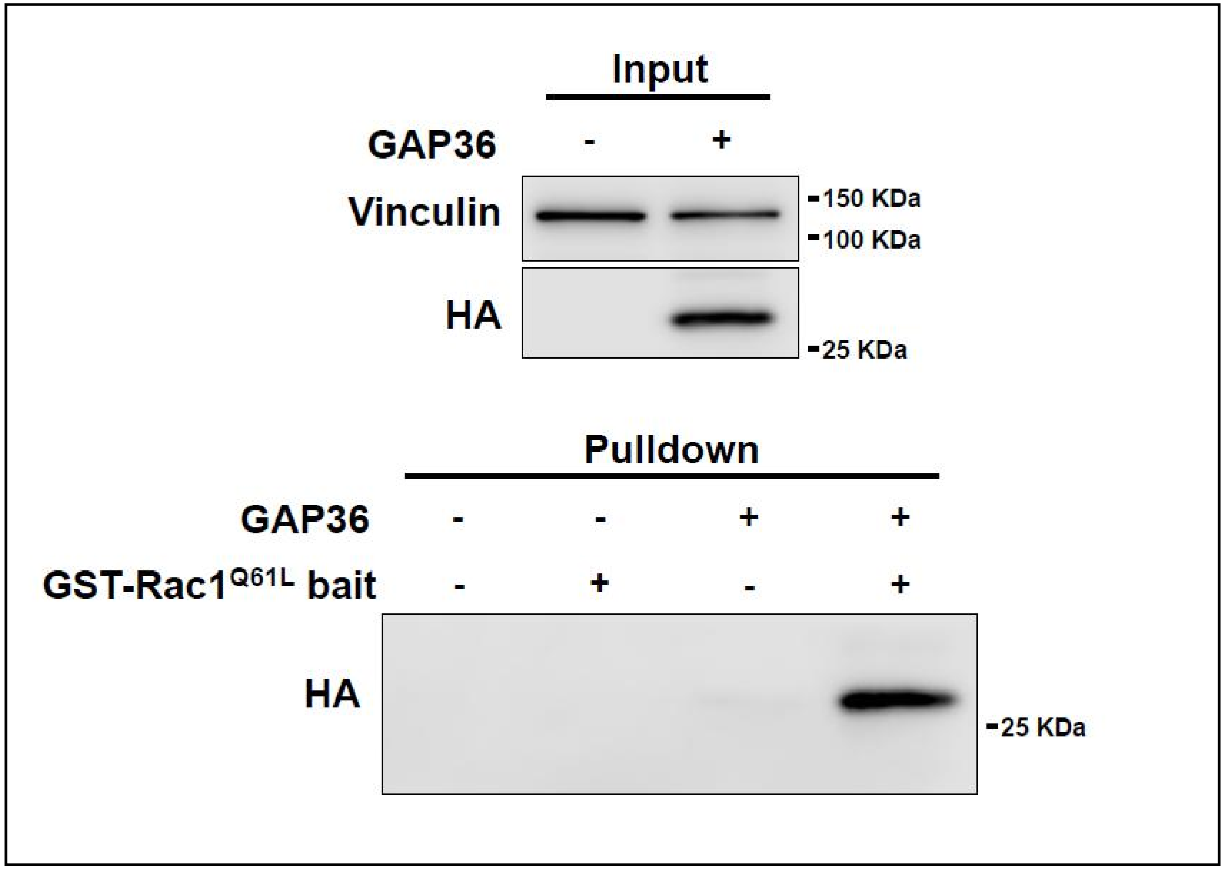

**Figure S6: ARHGAP36 interacts with RAC1.**

Pulldown assays using a clone of the GAP domain of ARHGAP36 and a recombinant constitutively active RAC1 mutant. The FLAG-HA tagged GAP domain of Arhgap36 (GAP36) was ectopically expressed in HEK293T cells and precipitated using a recombinant GST tagged active Rac1Q61L mutant. The top panel shows the input of control or FLAG-HA-GAP36 expressing cells and the bottom panel shows the precipitated GAP36, detected with anti-HA.

### SUPPLEMENTARY TABLES

#### Table S1. Putative loss-of-function variants of *ACTRT1* in public databases

| position | dnSNP | Major Allele | Minor Allele | Frequency# | Consequence | HGVS |
|---|---|---|---|---|---|---|
| ChrX:127185122 | rs1569354431 | T | - | 0.000005 (1/183050) | Frameshift | NM_138289.4:c.1064del, NP_612146.1:p.Gln355fs |
| ChrX:127185072 | rs867661986 | G | A | NA | Stop_gained | NM_138289.4:c.1114C>T, NP_612146.1:p.Gln372Ter |
| ChrX:127185181 | rs768201050 | - | ATCA | 0.000016 (3/183193) | Stop_gained | NM_138289.4:c.1002_1005dup, NP_612146.1:p.Arg336Ter |
| ChrX:127185201 | rs766490344 | T | A | 0.000005 (1/183246) | Stop_gained | NM_138289.4:c.985A>T, NP_612146.1:p.Lys329Ter |
| ChrX:127185247 | rs773801438 | C | - | 0.000005 (1/183215) | Frameshift | NM_138289.4:c.939del, NP_612146.1:p.Arg313fs |
| ChrX:127185329 | rs1223259859 | - | CTTCA | 0.000008 (1/125568)^ | Stop_gained | NM_138289.4:c.853_857dup, NP_612146.1:p.Cys286Ter |
| ChrX:127185462 | rs747807748 | TGT | T | 0.000005 (1/183135) | Stop_gained | NM_138289.4:c.723_724del, NP_612146.1:p.Tyr241Ter |
| ChrX:127185490 | rs1174530093 | C | - | 0.000005 (1/183221) | Frameshift | NM_138289.4:c.696del, NP_612146.1:p.Ser233fs |
| ChrX:127185564 | rs771674435 | GAGT | - | 0.000119 (15/125568)^ | Frameshift | NM_138289.4:c.619_622del, NP_612146.1:p.Leu207fs |
| ChrX:127185571 | rs1397178516 | G | T | 0.000005 (1/183275) | Stop_gained | NM_138289.4:c.615C>A, NP_612146.1:p.Cys205Ter |
| ChrX:127185639 | rs771087307 | - | T | 0.001822(373/204684) | Frameshift | NM_138289.4:c.547dup, NP_612146.1:p.Met183fs |
| ChrX:127185642 | rs760509819 | AG | - | 0.000024 (3/125568)^ | Frameshift | NM_138289.4:c.541_542CT, NP_612146.1:p.Cys182fs |
| ChrX:127185831 | rs767426435 | G | A | 0.000011 (2/183183) | Stop_gained | NM_138289.4:c.355C>T, NP_612146.1:p.Arg119Ter |
| ChrX:127185840 | rs776692535 | T | - | 0.00004 (1/183206) | Frameshift | NM_138289.4:c.346del, NP_612146.1:p.Arg116fs |
| ChrX:127185859 | rs1569354678 | G | - | 0.000005 (1/183222) | Frameshift | NM_138289.4:c.327del, NP_612146.1:p.Glu110fs |
| ChrX:127185864 | rs759455373 | - | T | 0.000005 (1/183243) | Frameshift | NM_138289.4:c.322dup, NP_612146.1:p.Met108fs |
| ChrX:127185877 | rs764199744 | TT | - | 0.000005 (1/183229) | Frameshift | NM_138289.4:c.308_309del, NP_612146.1:p.Gln103fs |
| ChrX:127185883 | rs1556035539 | - | G | 0.00108 (11/10195)* | Frameshift | NM_138289.4:c.303dup, NP_612146.1:p.Ser102fs |
| ChrX:127185907 | rs761972772 | AA | - | 0.000027 (5/183180) | Stop_gained | NM_138289.4:c.278_279del, NP_612146.1:p.Leu92_Phe93insTer |
| ChrX:127185909 | rs767686485 | GA | - | 0.000005 (1/183194) | Frameshift | NM_138289.4:c.274_275CT[1], |

| | | | | | | |
|---|---|---|---|---|---|---|
| | | | | | | NP_612146.1:p.Leu92_Phe93insTer |
| ChrX:127185911 | rs750649097 | A | - | 0.000005 (1/183181) | Frameshift | NM_138289.4:c.275del, NP_612146.1:p.Leu92fs |
| ChrX:127185918 | rs759293030 | T | A | 0.000005 (1/183169) | Stop_gained | NM_138289.4:c.268A>T, NP_612146.1:p.Lys90Ter |
| ChrX:127186142 | rs754366510 | T | - | 0.000011 (2/176668) | Frameshift | NM_138289.4:c.44del, NP_612146.1:p.Asp15fs |

### Frequency data was from GnomAD_exome if not marked. *: GO-ESP, NHLBI Grand Opportunity Exome Sequencing Project (ESP). ^: TOPMED, NHLBI Trans-Omics for Precision Medicine WGS.

**Table S2. Microdeletions encompassing *ACTRT1* in DECIPHER and associated clinical phenotypes.**

| Location | Genes | Size | Inheritance / Genotype | Pathogenicity | Phenotypes |
|---|---|---|---|---|---|
| ChrX:127184000-127358000 | *ACTRT1* | 174Kb | Maternally inherited/Heterozygous | Uncertain | Not available |
| ChrX:126958431-127369766 | *ACTRT1* | 411.34Kb | Unknown/Heterozygous | Unspecified | Atrial septal defect, Abnormality of the outer ear, Blepharophimosis, et al. |
| ChrX:126779219-127441765 | *ACTRT1* | 662.55Kb | Inherited from normal patient/Hemizygous | Unspecified | Abnormality of cardiovascular system morphology |
| ChrX:126779219-127441765 | *ACTRT1* | 662.55Kb | Unknown/Hemizygous | Unspecified | Proptosis, Neonatal hypotonia, Delayed gross motor development |
| ChrX:126117880-127358432 | *ACTRT1* | 1.24Mb | Inherited from normal patient/Heterozygous | Unspecified | Deeply set eye, Hypertelorism, Epicanthus, et al. |
| ChrX:124439330-127799936 | 7 genes | 3.36Mb | Inherited from normal patient/Hemizygous | Unspecified | Truncal obesity, Abnormal eyelid morphology, Macrodontia, et al. |

#### Table S3. Primers used for qPCR and long-range gap-PCR in BDCS families.

| Family | name | sequence | location |
|---|---|---|---|
| F1 | XLB4F | CAGACACCAGCTGTGCAATGTA | chrX:130280109-130280257 |
| | XLB4R | TGGTCCTTGGATCCCATAGC | |
| | F1LB2-1F | CATTGAGCCATGCACCTTGT | chrX:130285223-130285348 |
| | F1LB2-1R | CAATTGGTGCGCAAGACACT | |
| | F1LB2-2F | TCCATGCCAATAGAATGCTGTT | chrX:130292585-130292699 |
| | F1LB2-2R | AATGCTCACCCAAATCACAGAA | |
| | F1LB2-3F | ATGGCTCTGACAGGGAATGACT | chrX:130298950-130299060 |
| | F1LB2-3R | GCAGGACCACAATGTTTTGGA | |
| | XQMF | TGCCTCCAGTAGTCCATTGACA | chrX:130348186-130348315 |
| | XQMR | CCAAACCTCATCCAGAGTGCTT | |
| | RB3-1F | TCAGACATTTTCATTTGCCAGTCTT | chrX:130378118-130378232 |
| | RB3-1R | TGACCAGCAAACAGGAAATGC | |
| | RB3-3F | TGGAAAAATCAAAGAGTGAAAATCCT | chrX:130386198-130386290 |
| | RB3-3R | TGCGCTTCGTTGTTGATTACA | |
| F3, F5, F8 | XQMF | TGCCTCCAGTAGTCCATTGACA | chrX:130348186-130348315 |
| | XQMR | CCAAACCTCATCCAGAGTGCTT | |
| F2 | F2LB2-1F | TCCGAGATTCTTTTGACCCACTA | chrX:130233886-130233993 |
| | F2LB2-1R | CAGCAGAGCTGCAATATTACTGAAA | |
| | F2LB2-2F | TTTCTAAAGCGTTGGCACCAT | chrX:130252673-130252818 |
| | F2LB2-2R | TCAGACCTGCCAGTACGGTTT | |
| | F2LB2-3F | CACTGAATTCCTTTGGTGACTTTG | chrX:130253436-130253546 |
| | F2LB2-3R | GCAACATTGGTTTGATGAAAGAGA | |
| | XQMF | TGCCTCCAGTAGTCCATTGACA | chrX:130348186-130348315 |
| | XQMR | CCAAACCTCATCCAGAGTGCTT | |
| | RB3-1F | TCAGACATTTTCATTTGCCAGTCTT | chrX:130378118-130378232 |
| | RB3-1R | TGACCAGCAAACAGGAAATGC | |
| | RB3-3F | TGGAAAAATCAAAGAGTGAAAATCCT | chrX:130386198-130386290 |
| | RB3-3R | TGCGCTTCGTTGTTGATTACA | |
| F4 | F4B1-9F | CAGGCTGCTATGGTATCAAATTTG | chrX:130338549-130338647 |
| | F4B1-9R | ATCTGAGCAGGAGAGCTTATGCA | |
| | F4B1-10F | ACTCCTGGGATAGGGCCAGAT | chrX:130342136-130342225 |
| | F4B1-10R | CTTGACAGCCACTGCCATATTG | |
| | XQ-F1 | TGGTGTTAATTGGTCGCATCTG | chrX:130347208-130347297 |
| | XQ-R1 | AATAACTCGCCCTATGCCCATCT | |
| | XQ-F2 | TCTCACCATTAGGGATGGACTGT | chrX:130352328-130352417 |
| | XQ-R2 | CCTGCCGATCATCTCACAAA | |

| | | | |
|---|---|---|---|
| | XQ-F3 | GTGGTGCCCAAAGACCCATA | chrX:130356319-130356408 |
| | XQ-R3 | GGTCACAGGGCAGACTGATCTC | |
| | F4RF1 | TGGCCCCAACGTTCCTAGA | chrX:130357253-130357359 |
| | F4RR1 | TCCCTCATAGCATTGATCACAATAG | |
| | F4B1-12F | TTCTATGCCTGCTCCATTCC | chrX:130360154-130360250 |
| | F4B1-12R | GGGCTAGCACCATGAGGGTAT | |
| | XQ-F4 | TTATCACAGGTTCCAGATGGAGAA | chrX:130363861-130363951 |
| | XQ-R4 | TGATGAAAACCACCCTCCTATCTAC | |
| F6 | F4B1-9F | CAGGCTGCTATGGTATCAAATTTG | chrX:130338549-130338647 |
| | F4B1-9R | ATCTGAGCAGGAGAGCTTATGCA | |
| | F4B1-10F | ACTCCTGGGATAGGGCCAGAT | chrX:130342136-130342225 |
| | F4B1-10R | CTTGACAGCCACTGCCATATTG | |
| | F4B1-12F | TTCTATGCCTGCTCCATTCC | chrX:130360154-130360250 |
| | F4B1-12R | GGGCTAGCACCATGAGGGTAT | |
| | F4B1-13F | TGATGCTTCCCCCACAAAAGA | ChrX:130362889-130362981 |
| | F4B1-13R | TCATTCTGTGATCCCCCACAT | |
| F7 | F4B1-6F | TTGATTCCAAGTTGGTTACCCCATA | chrX:130325368-130325457 |
| | F4B1-6R | TGGCTTTGAAGCATGGGAAA | |
| | F7B1-1F | AGGTATTGTCATTGGCCCACAT | chrX:130327565-130327657 |
| | F7B1-1R | GCATGAAGTTGCCTTTAAGATCAG | |
| | F4B1-7F | CAGGGCCATAGGTGGATCAT | chrX:130329905-130329994 |
| | F4B1-7R | CAAATAACATTCAGCCCAGTTTCC | |
| | F4B1-8F | CCCTATAGCAAGCCCTGTTCTAAA | chrX:130333001-130333092 |
| | F4B1-8R | GGCCTCCAGATACAATCCATAATTA | |
| | F4B1-9F | CAGGCTGCTATGGTATCAAATTTG | chrX:130338549-130338647 |
| | F4B1-9R | ATCTGAGCAGGAGAGCTTATGCA | |
| | RB3-1F | TCAGACATTTTCATTTGCCAGTCTT | chrX:130378118-130378232 |
| | RB3-1R | TGACCAGCAAACAGGAAATGC | |
| | RB3-3F | TGGAAAAATCAAAGAGTGAAAATCCT | chrX:130386198-130386290 |
| | RB3-3R | TGCGCTTCGTTGTTGATTACA | |
| **For Long-range gap-PCR** | | | |
| **Family** | **name** | **sequence** | |
| F1，F3，F5 and F8 | RB3-1F | TCAGACATTTTCATTTGCCAGTCTT | |
| | F1LB2-2R | AATGCTCACCCAAATCACAGAA | |
| F2 | RB3-1F | TCAGACATTTTCATTTGCCAGTCTT | |
| | F2LB3-4R | TGCCTGAGATAGCCATAATAGAAATC | |
| F4 | F4B1-12F | TTCTATGCCTGCTCCATTCC | |
| | F4B1-10R | CTTGACAGCCACTGCCATATTG | |
| F6 | F4B1-10R | CTTGACAGCCACTGCCATATTG | |
| | F4B1-12F | TTCTATGCCTGCTCCATTCC | |
| F7 | RB3-1F | TCAGACATTTTCATTTGCCAGTCTT | |

| | | |
|---|---|---|
| | F7B1-1R | GCATGAAGTTGCCTTTAAGATCAG |
| Primers located outside the duplication region are colored in red. | | |
